# Comprehensive Review of Adverse Effects in Minimally Invasive Cosmetic Procedures: Insights from a Decade of Clinical Data

**DOI:** 10.1101/2024.08.23.24312506

**Authors:** Cristina Guadalupe Beltrán Muñoz, Elissa Bracamontes Perez, Fernanda Corrales Bay, Aimee Adriana Barajas Nieto, Kate Georgette Rojo Navarro

## Abstract

**Introduction:** Minimally invasive cosmetic procedures are medical interventions that do not involve the use of surgical incisions. They are usually based on the administration of biocompatible compounds into subcutaneous or dermal tissue. Although considered low-risk, they can have significant adverse effects such as tissue damage or occlusion of superficial blood vessels. Describing their complications is necessary to ensure the responsible use of these resources.

**Objective:** To describe the adverse effects secondary to the use of minimally invasive cosmetic procedures.

**Materials and Methods:** Studies describing the adverse effects of minimally invasive cosmetic procedures were searched in PubMed, Scopus, Embase, and Google Scholar from January 2012 to December 2023.

**Results:** A total of 98 articles were included. The administration of hyaluronic acid, botulinum toxin, silicone, and autologous fat grafts were the most commonly used procedures. Local symptoms were the most commonly reported reactions (53 studies). Meanwhile, temporary or permanent blindness was reported in 10 studies, immunological alterations in 8, severe pulmonary involvement in 6, neurological damage in 5, arterial occlusion in 3, and 2 studies reported fatalities.

**Conclusion:** Minimally invasive cosmetic procedures can cause local complications in the majority of cases, but they can also lead to fatal outcomes, particularly in unprofessional settings. Undergoing these techniques requires a comprehensive professional assessment and close follow-up by licensed personnel.

## INTRODUCTION

Currently, there is a significant and growing demand for medical procedures aimed at cosmetic enhancement^1,2^. This trend is particularly evident in the increasing popularity of minimally invasive procedures, which are favored for their effectiveness, ease of application, and shorter recovery times^3,4^. These procedures often involve the administration of various substances, such as hyaluronic acid, botulinum toxin, autologous fat grafts, and other cellular products, among others^5–8^. While these compounds have been proven to be stable, biocompatible^9^, biodegradable and non-toxic^10,11^, the detailed mechanisms underlying their cellular effects, metabolism, and excretion remain partially understood^12^. This uncertainty raises concerns about potential tissue damage, particularly across different skin layers and superficial neurovascular bundles^13^. Despite the widespread use of these procedures, most research has predominantly focused on documenting the adverse effects associated with the most commonly performed interventions^14^, or those linked to specific application techniques^15,16^. Furthermore, the unlicensed practice of cosmetic procedures is alarmingly common, which exacerbates the potential risks these interventions present^17^. This leaves a gap in understanding the broader spectrum of risks these procedures may pose. Therefore, we conducted a comprehensive review of the literature with the objective of describing the complications experienced by patients undergoing minimally invasive cosmetic procedures over the past decade. Additionally, we aim to provide a broader perspective on the potential risks associated with these interventions, emphasizing the importance of their responsible use.

## METHODS

Using the PICO strategy, we conducted a systematic search in the PubMed, Embase, Scopus and Google Scholar repositories, with the following keywords: “*aesthetic medicine and side effects*” OR “*cosmetic medicine and side effects*” OR “*minimally invasive procedures and iatrogenic*” OR “*filler injection and side effects*” OR “*minimally invasive aesthetic medicine and adverse outcomes*”. Only original articles published between January 2012 and December 2023, written in English or Spanish, and involving human participants undergoing minimally invasive cosmetic procedures (e.g., appearance modification, enhancement of attractiveness, etc.) were included”. Systematic reviews and meta-analyses, expert consensus, pre-clinical experimental studies and in-vitro studies were excluded, as well as studies with unavailable content or imprecise information regarding the interventions used. Articles were filtered by title and abstract by five reviewers, and those with relevant information were selected by consensus. Information on the type of procedure, occurrence of adverse events, and other covariates or interest, was collected for all selected articles.

## RESULTS

We encountered a total of 238 candidate articles, of which 55 were excluded for containing information unrelated to the aims of this review, 18 for being in a different language, 10 for being duplicates and 57 due to incomplete data, yielding a total of 108 eligible articles. Of the included articles, a total of 33 studies were randomized and 5 were nonrandomized clinical trials, 8 were retrospective cohort studies and 3 were prospective cohort studies. Additionally, we also included 10 case series, 37 case reports, and 2 “*brief report*s” (**Figure 1, Table 1**).

**Figure 1.**
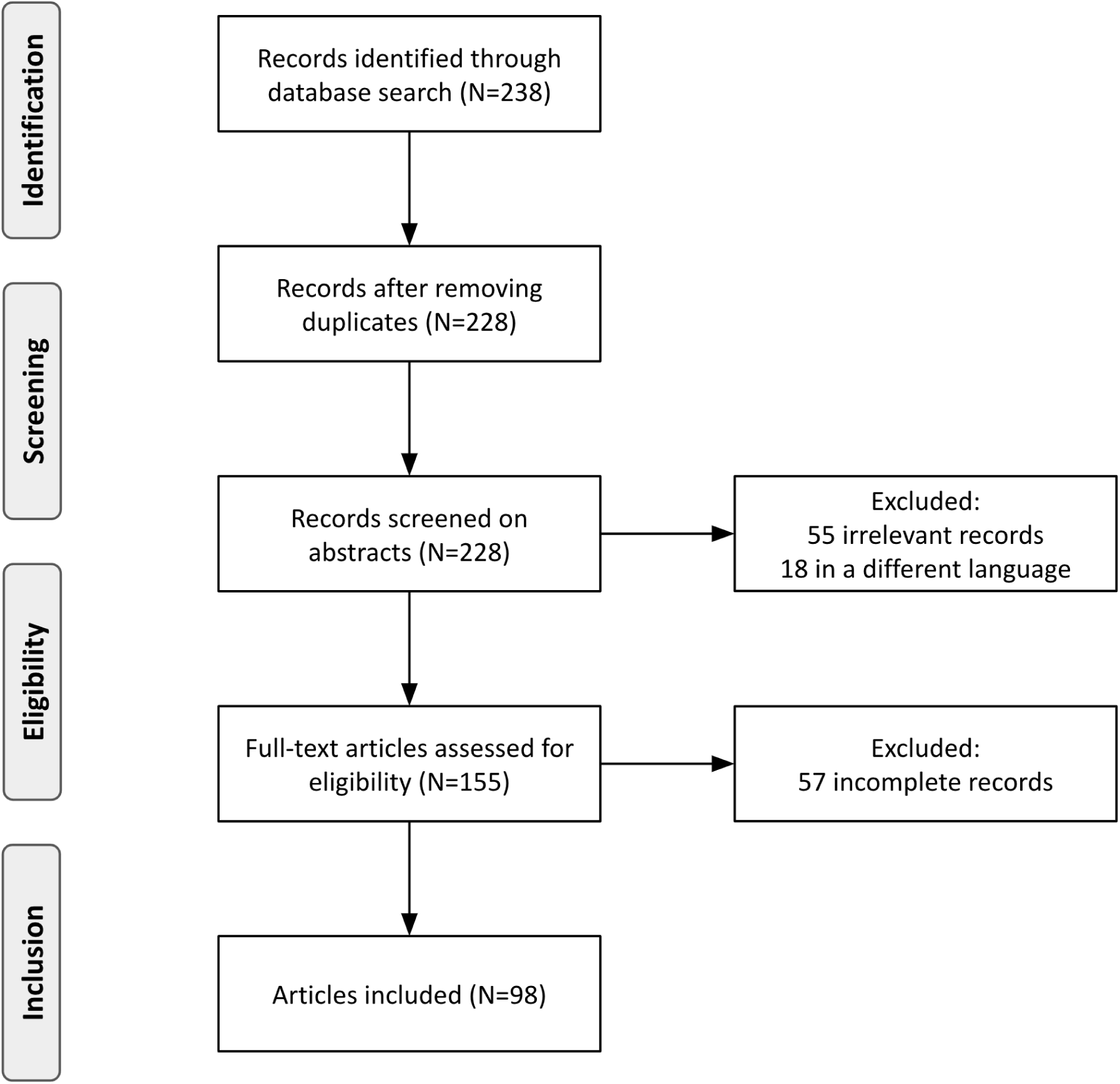
Summary of research strategy.

**Table 1.**
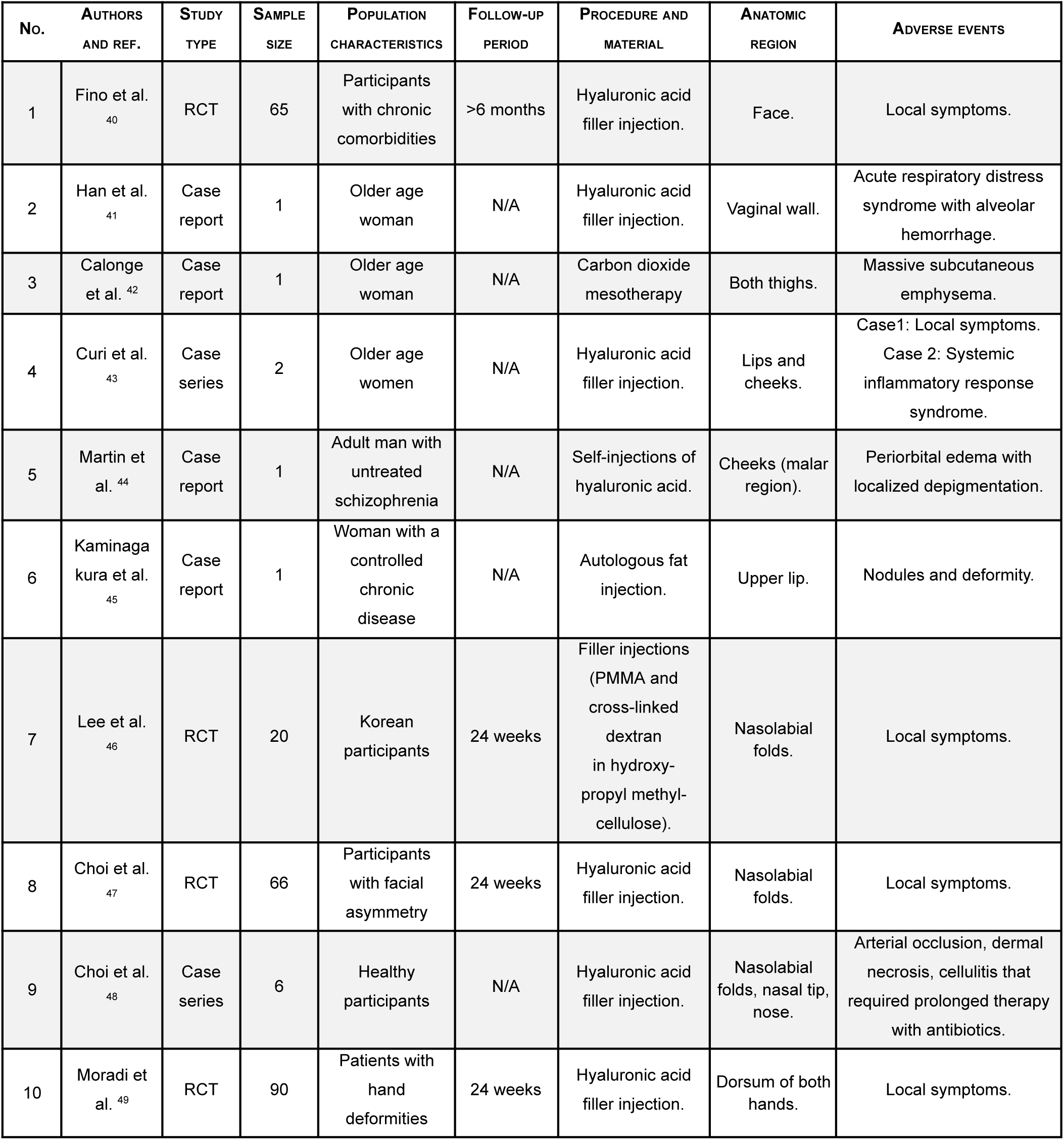
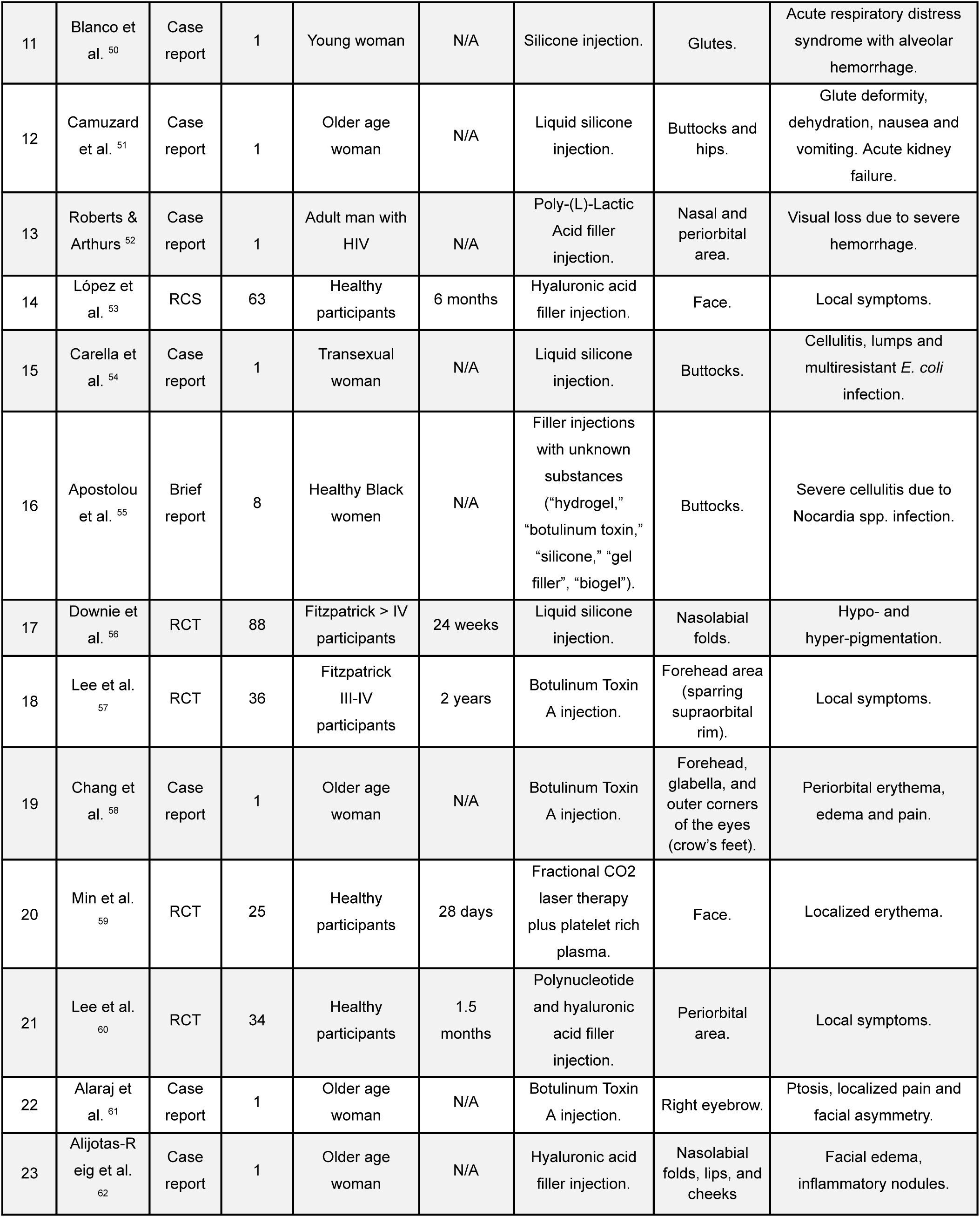
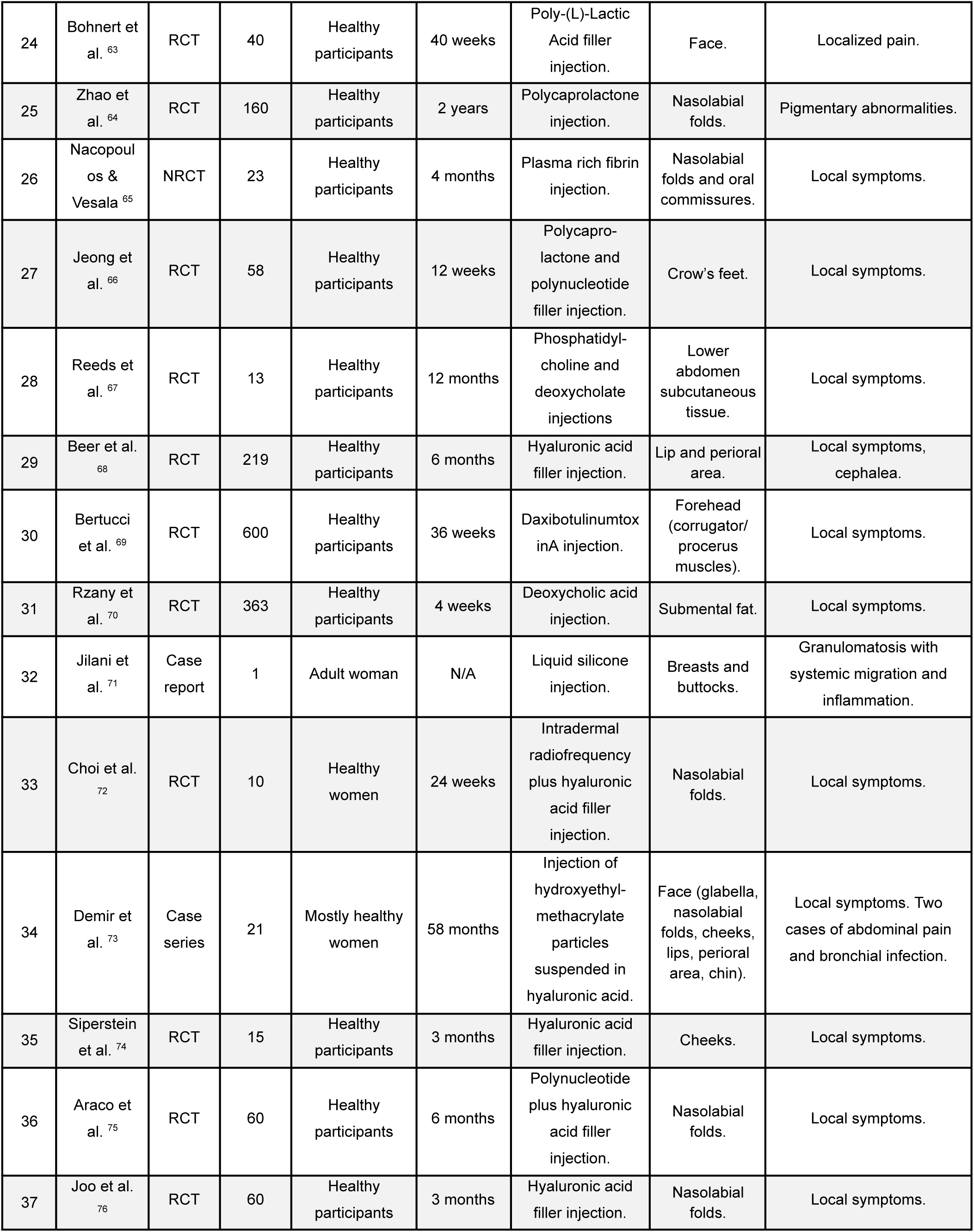
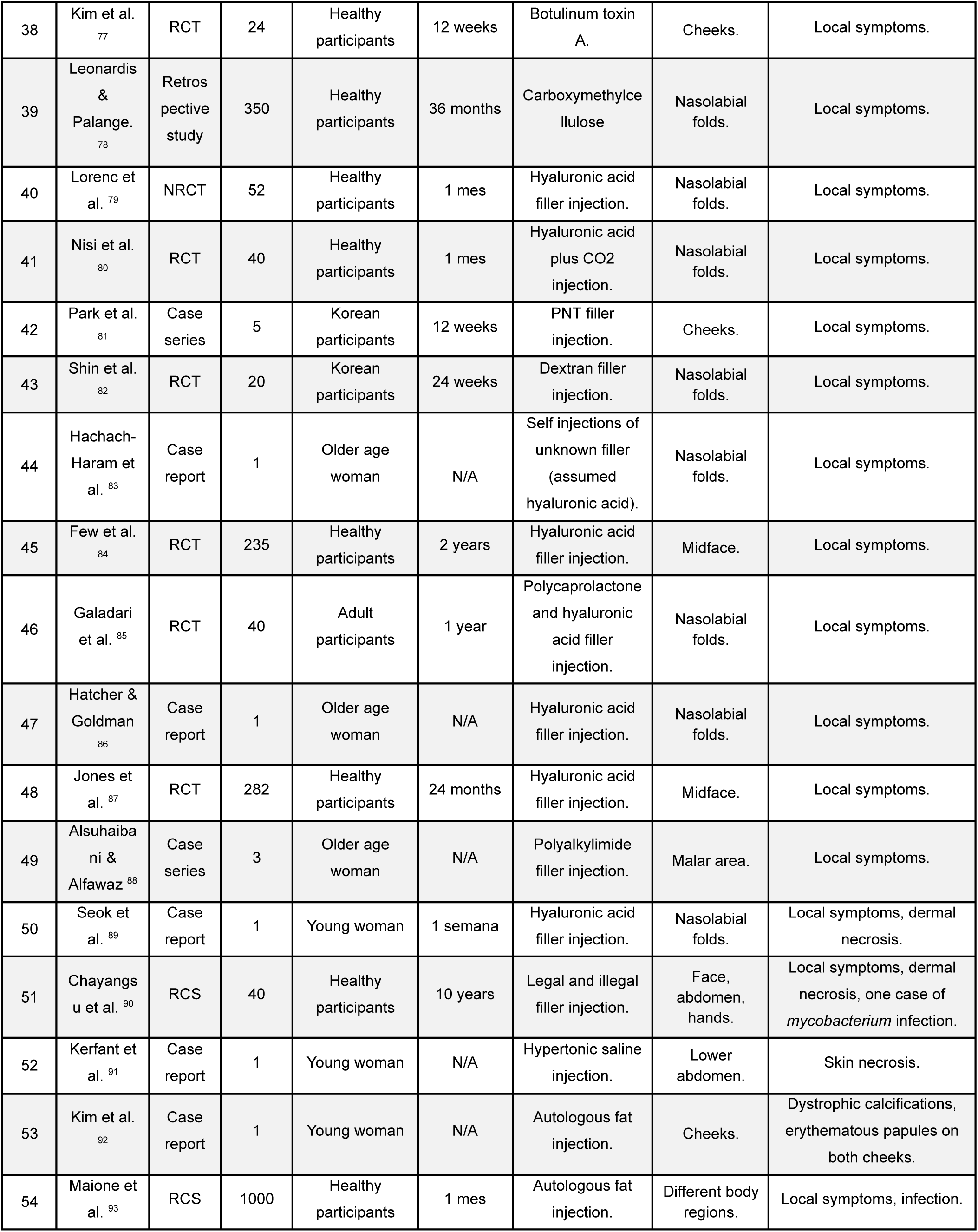
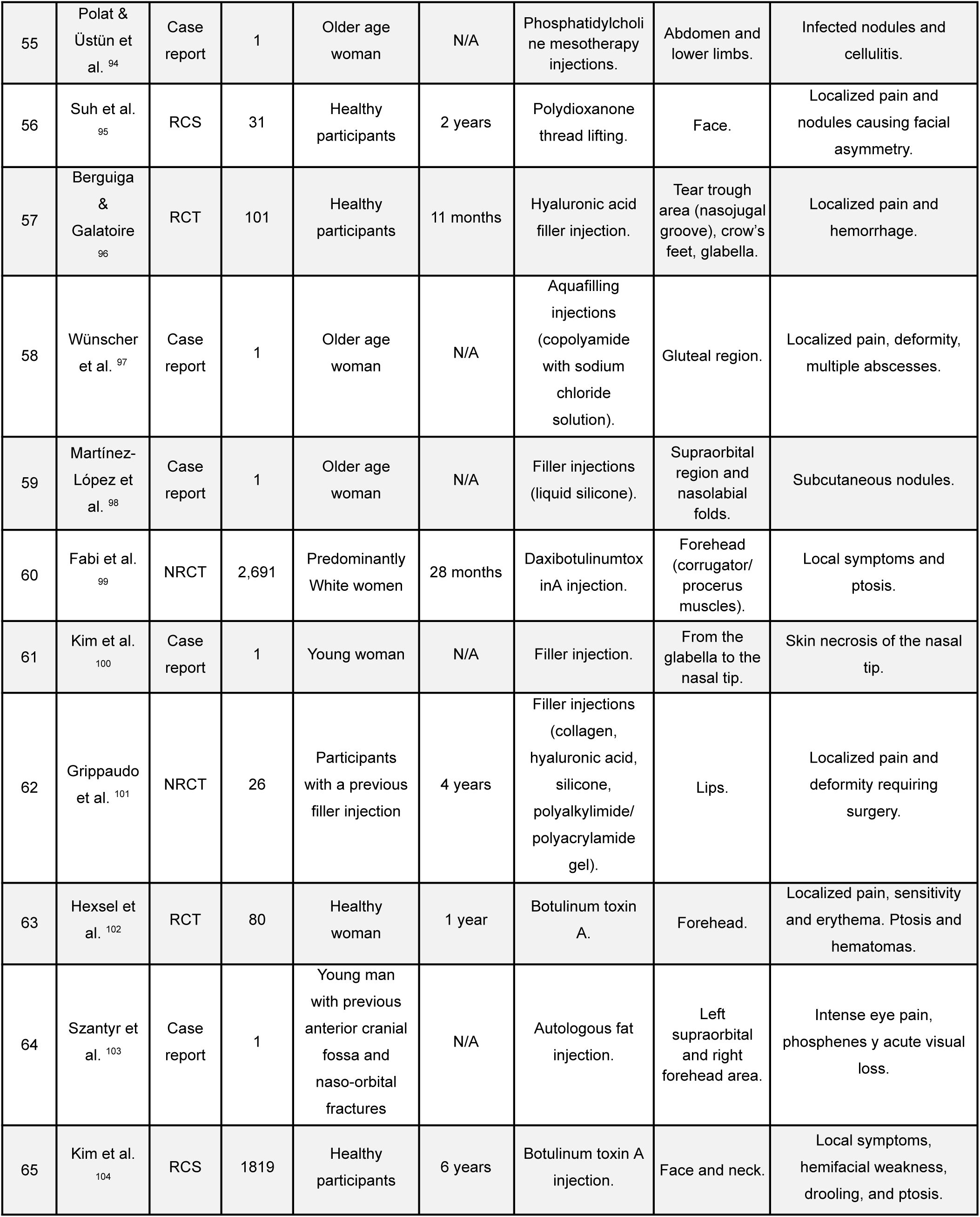
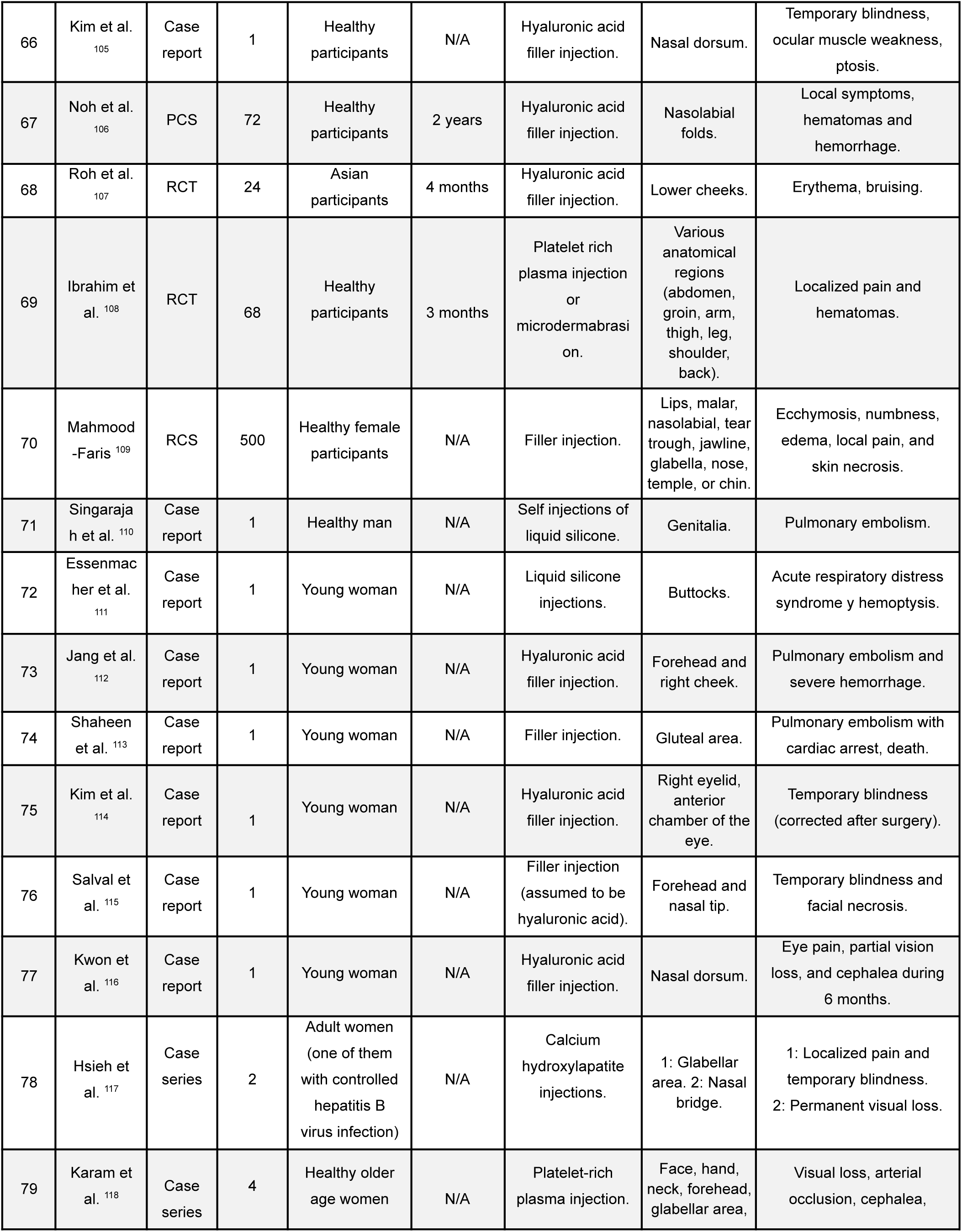
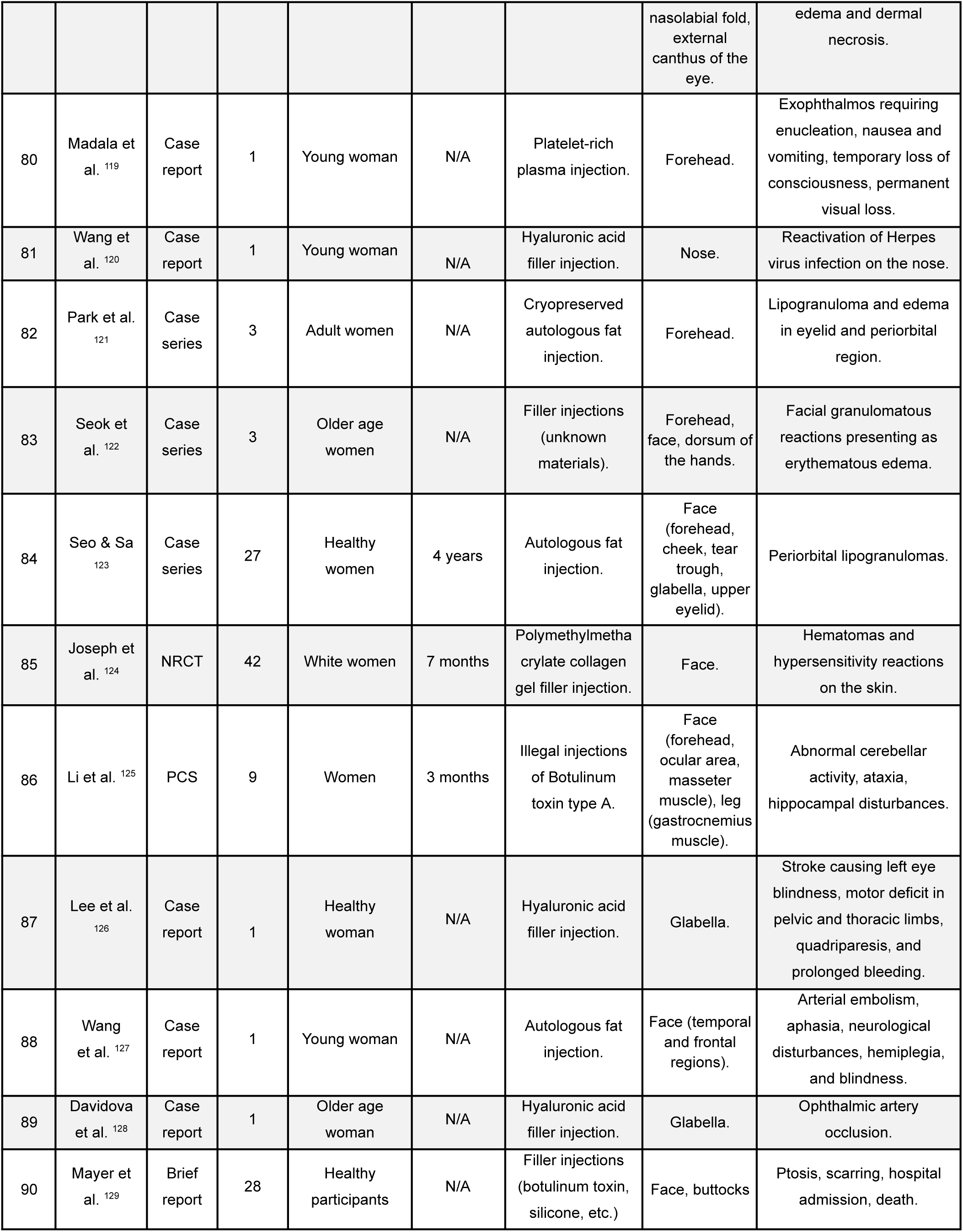
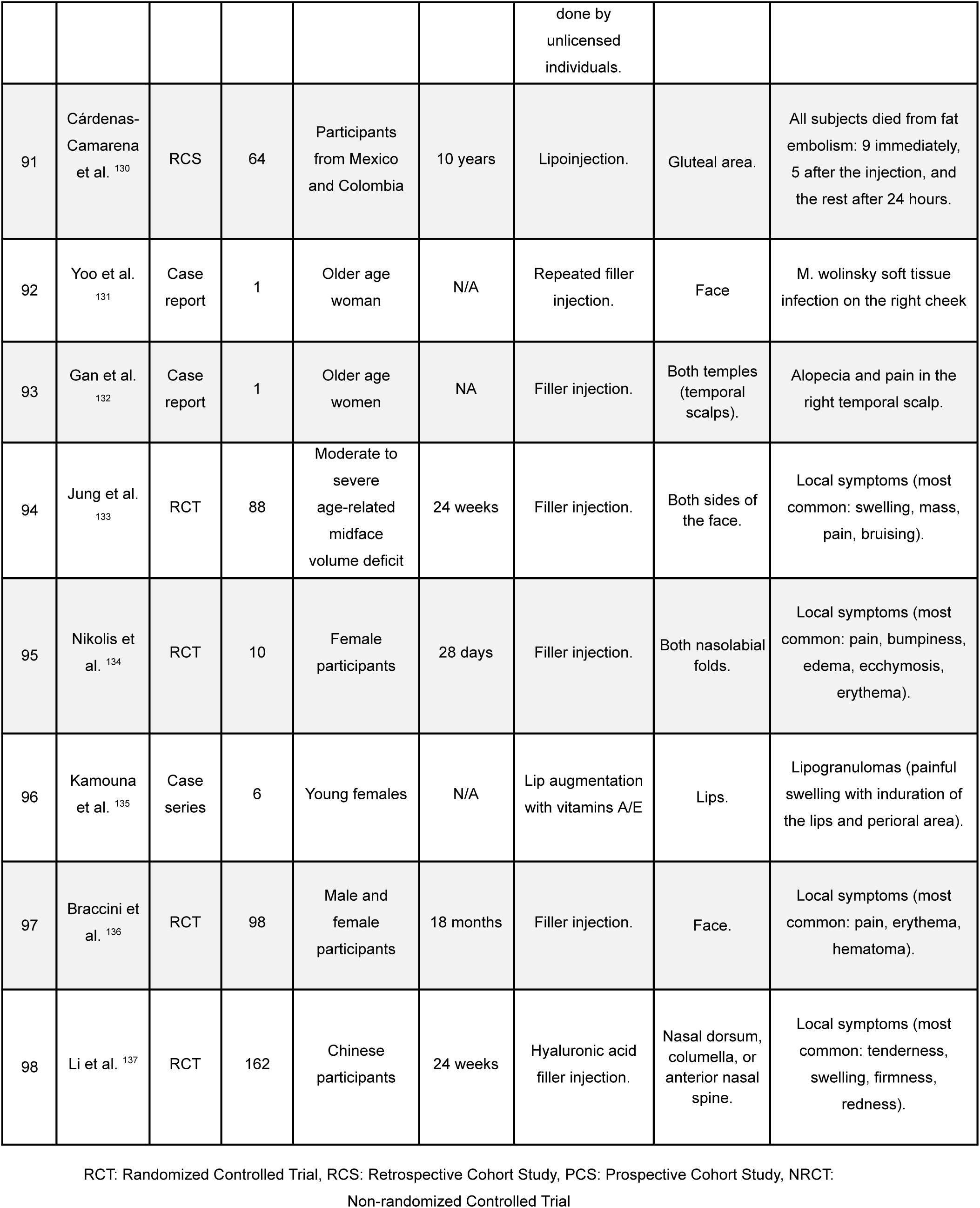
Summary of Analyzed Resources.

### General characteristics of participants

The total sum of the population analyzed in the studies amounted to 10,231 patients. All participants were adults, aged between 18 and 80 years, with the majority being cisgender women in their third to sixth decade of life. Overall, most of the population was healthy and all of them had voluntarily undergone the procedures. Only participants from 17 studies presented with a relevant medical history, including participants with chronic degenerative diseases (9 studies), individuals with facial and/or body malformations (2 studies), with active infections including HIV and COVID-19 (4 studies), transgender participants (1 study), and a patient with a history of autoimmune disease (1 study) (**Table 1**). Additionally, the race/ethnicity of most participants was White, followed by Korean and Mestizo populations. Only 3 studies evaluated the effects of these interventions on dark-skinned populations (Fitzpatrick classification > IV).

### Procedures performed

Most studies focused on the administration of biocompatible substances of various types into superficial anatomical planes of the subjects to model body structures and enhance the attractiveness of the treated area. The most frequently used compound was commercially formulated hyaluronic acid, utilized in more than 40 studies. Additionally, over half of these studies used combinations of this substance with solvents, local anesthetics (mainly lidocaine), oils, or other components. In 3 of these 62 studies, the origin and composition of the administered hyaluronic acid could not be identified. There was also a noteworthy use of botulinum toxin, administered in 10 studies either as a standalone component or in combination with other substances. Liquid injectable silicone was used in 7 studies. The use of autologous cell products was also prevalent, such as fat grafts (7 studies), platelet-rich plasma (4 studies) or plasma-rich fibrin (1 study). Other substances used less frequently included carbon dioxide, polynucleotide, dextran, phosphatidylcholine, etc.

### Intervened areas and responsible health personnel

The most frequently intervened anatomical area was that of the face, chin, and lips (79 studies), particularly in nasolabial folds, cheek areas, and other various sites across the face. This was followed by the gluteal region (16 studies), periorbital, nose and glabella region (8 studies), abdomen (6 studies), and genitalia (2 studies). In most records, the interventions were performed by a specialist physician trained in plastic surgery or another unspecified field. However, there were several instances where the interventions were carried out by unqualified personnel, including uncertified clinics or physicians without a license to perform these procedures. In 3 studies, the intervention was self-administered.

### Follow-up of subjects

Excluding case reports and case series studies, the follow-up period for subjects within the studies was highly variable. The follow-up period was less than 6 weeks in 7 studies, between 6 weeks and 6 months in 10 studies, and between 6 to 12 months in 20 studies. Additionally, 8 studies had a duration of around 2 years, 3 a duration of 2-5 years, and 3 had a follow-up period exceeding 5 years.

### Minor and local adverse effects

All studies followed common guidelines for recording adverse effects following cosmetic procedures in humans and were classified in early and late complications. Most of the adverse effects observed were of low severity and appeared immediately or within a few days of the intervention. In the majority of these studies, only minimal discomforts such as periorbital edema, erythema, and localized pain were found. Three studies recorded skin pigmentation alterations with patches of hypo- and hyperpigmentation. In addition to the described discomforts, thirty other studies found cases of swelling, more severe edema, and deformities, with all symptoms disappearing within 48 hours to 7 days. Additionally, eleven studies reported the presence of lumps, nodules, deformities with “unaesthetic” results, infections, cellulitis of varying intensity, and dermal necrosis with prolonged or permanent duration in subjects who underwent these interventions. On the other hand, seven studies reported additional complications such as eyelid ptosis and/or extraocular muscle weakness. Episodes of hematoma and mild hemorrhage were reported in five studies.

### Severe adverse effects

The most significant adverse effects were recorded in case report studies, populations with medical history of comorbidities, or studies with over 5 years of follow-up. A total of seven cases reported severe pulmonary involvement due to embolisms that caused acute respiratory distress syndrome, pulmonary hemorrhage, and/or alveolar hemorrhage. Moreover, eight studies recorded loss of visual acuity, temporary blindness, or permanent unilateral vision loss. Another eight studies presented immunological alterations such as granuloma formation, autoimmune syndromes, and reactivation of a herpes simplex virus infection. Five studies reported severe neurological symptoms, such as ataxia, vertigo, paralysis of the thoracic limbs, facial paralysis, etc., likely due to damage to the cerebellar region, hippocampus, and temporal cortex. Three papers reported notable cases of arterial occlusion; one causing severe necrosis and infection after hyaluronic acid injections on the nose and nasolabial folds, requiring prolonged treatment with antibiotics; the second generating visual loss due to ophthalmic artery obstruction after platelet-rich plasma injections into the face; and the third resulting in unilateral blindness after hyaluronic acid injections in the glabellar area. One case of kidney failure was reported in a participant after undergoing liquid silicone injection in the buttocks and hips. One study reported 4 hospitalizations and 8 deaths in a cohort of 28 patients who underwent filler injections done by unlicensed individuals. Finally, a report on Mexican and Colombian participants documented 64 deaths, 9 of which occurred immediately after lipo injections in the gluteal area. **Table 2** shows the summary of the most common adverse effects found in the population and their approximate incidence per 1000 interventions performed sorted in descending order.

**Table 2.** Incidence of Adverse Events per 1000 Procedures Performed in the Population.

| **Adverse event** | **Occurrences per 1000 Procedures** |
| --- | --- |
| Local complications (pain, erythema, edema, temporary deformity, hypersensitivity) | 363.308 |
| Subcutaneous hemorrhage, hematomas | 55.127 |
| Asymmetry due to nodules, lumps, or calcifications, with non-aesthetic results | 28.443 |
| Pigmentary disturbances | 12.022 |
| Fatal fat embolism | 7.135 |
| Subocclusion or occlusion of the ophthalmic artery | 5.865 |
| Blepharoptosis | 5.376 |
| Temporal blindness | 3.03 |
| Cellulitis | 1.173 |
| Dermal and annex necrosis | 0.977 |
| Permanent blindness | 0.684 |
| Cerebral infarction | 0.489 |
| Acute respiratory distress syndrome | 0.489 |
| Pulmonary hemorrhage | 0.391 |
| Mycobacterial infection | 0.391 |
| Pulmonary embolism | 0.293 |
| Severe autoimmune reaction | 0.293 |
| Severe bronchial infection | 0.195 |
| Reactivation of herpes virus infection | 0.098 |
| Livedo Racemosa | 0.098 |
| Acute kidney injury | 0.098 |

The adverse effects presented correspond to the total number of interventions performed, without specific distinction of the procedure or technique implemented.

## DISCUSSION

In this work, we found various types of adverse effects reported in the medical literature after minimally invasive cosmetic procedures. Although the majority of recorded events corresponded to mild and localized complications with a rapid resolution, the presence of more severe and sometimes fatal complications pose a significant potential risk and calls for a responsible use of these procedures. The studies that reported the longest-lasting adverse effects were largely retrospective studies and clinical trials with follow-up periods extending beyond six months. This longer duration is critical for identifying late-onset complications that may not appear within the shorter follow-up periods commonly used in many studies. As a result, many of these complications may be underreported in the literature, since the delayed onset of these effects makes it difficult to establish a clear causal relationship between the procedure and the adverse outcomes^13,15^. This issue is especially significant when considering that short-term patient satisfaction, along with the heavy promotion of these procedures as safe and effective, might lead both patients and healthcare providers to downplay or overlook the potential for long-term adverse effects associated with cosmetic interventions. The focus on immediate results can create a false sense of security, where the benefits seem to outweigh any potential risks, thereby obscuring the true safety profile of these procedures^18^.

Another significant finding of this review is that in almost all studies, women represented the predominant population undergoing these procedures. While it is increasingly common for men to undergo aesthetic procedures, previous studies indicate that the perception of artificial enhancement of appearance differs by sex, with men preferring specific and more discrete procedures such as jawline contouring and hair implants^19^. Furthermore, a significant number of patients were Asian, particularly of Korean origin. Previous studies indicate the persistence of rigid gender roles in Korean society, which directly influence female self-esteem and personal well-being^20^, as well as the expected beauty standards in this population^21^, which likely has a large impact in their use of cosmetic procedures.

The complications described in this review can be grouped into two broad categories: those unrelated to ischemia (pain, erythema, granulomas, infections, etc.), and those secondary to ischemia (arterial occlusion, necrosis, etc.)^22^. The former were the most prevalent type of complication in the review, including aesthetic alterations such as the formation of nodules and facial asymmetries. The latter can be subdivided into acute or chronic adverse effects. In this review, chronic complications were the most severe, often requiring multidisciplinary and even surgical intervention, as has been previously described for such complications^23^. Factors associated with these complications are varied, including healthcare professional expertise^24^, presence of chronic comorbidities, substance abuse^25^, anatomical positioning of superficial blood vessels, and the molecular size of injected compounds^26^. However, most studies were conducted on a healthy population that voluntarily underwent the intervention, so there might be factors or pathological conditions unaccounted for during recruitment that favored the occurrence of long-term complications.

In this review, the most frequently administered substances were hyaluronic acid, botulinum toxin, fat implants, and silicone. Although ultra pure hyaluronic acid is primarily obtained through recombinant DNA technology, contaminants in the stabilizers used in its formulation can generate a delayed immune response that predisposes participants to fibrosis and granulomas^27^, as reported in many studies. Interestingly, the commercial formulations described with greater detail were the ones associated with milder adverse effects, a potential explanation is that different hyaluronic acid formulations are designed for specific skin layers^28^; thus, their correct choice and administration may reduce the risk of complications. Moreover, their incorrect use is particularly dangerous in certain highly vascularized anatomical areas, such as the glabella and the periocular regions, where small superficial manipulations can trigger emboli formation^29,30^. On the other hand, injections of botulinum toxin were reported in many studies in which granulomas, cellulitis, and other infections by atypical microorganisms (i.e., mycobacterium), were also present, which is consistent with previous reports^31,32^. Significant complications were also recorded following the use of silicone, such as implant migration and autoimmune reactions. This can be due to minor contaminations in these implants, in which the presence of microorganisms along with the slow degradation of silicone compounds can favor chronic immune reactions that only become apparent after long follow-up periods^33^. Lastly, most hospital admission and deaths recorded were secondary to severe complications associated with embolism with arterial obstruction and involvement at the pulmonary, cerebral, and retinal levels. These events can occur after administration of any compound, but they were more common following injections of autologous fat implants and some types of hyaluronic acid formulations. Although fat implants undergo comprehensive filtration processes^34^, migration through the bloodstream remains a significant risk associated with their application^35^. On the other hand, the higher viscosity and hydrophilicity of hyaluronic acid formulations facilitates its adhesion to vascular walls, increasing the risk of vascular-related complications^36^. Although it is believed that negative aspiration before injection may reduce risk, in reality complications can still occur due to a combination of factors such as perivascular injuries or anatomical positioning, which can favor the release of the administered compound into the bloodstream^37^. In this regard, a thorough knowledge of injection site anatomy and patient history (e.g., presence of chronic conditions and allergies, use of blood thinners), a correct injection technique, and the application of adequate antiseptic procedures with non-irritating substances are more reliable safety measures to prevent undesirable adverse effects^38^. Additionally, the use of ultrasound in these interventions should be considered as a safer approach, offering enhanced precision and potentially reducing the risk of complications.^39^.

In conclusion, minimally invasive aesthetic procedures, while popular, can result in adverse effects of varying severity. The possibility of unnoticed long-term effects poses an additional concern for both users and healthcare professionals. To minimize risks, it is highly recommended to reduce patient risk factors and enhance procedural accuracy through techniques such as ultrasound guidance. Finally, ensuring that these procedures are exclusively performed by qualified medical personnel is crucial to maintaining patient safety and the overall integrity of the field.

## Data Availability

The datasets used and/or analyzed during the current study are available from the corresponding author upon reasonable request.

## AUTHOR CONTRIBUTIONS

CGBM, EBP, FCB, AABN, and KGRN were jointly involved in selecting and reviewing the resources, as well as in drafting and structuring the manuscript. CGBM was responsible for the conception, integration, and final structuring of the manuscript.

## ETHICAL APPROVAL AND CONSENT TO PARTICIPATE

Not applicable. This study only uses non-identifiable patient information.

## CONSENT FOR PUBLICATION

Not applicable. The publication of these data does not compromise the anonymity or confidentiality of any patient.

